# Discriminatory ability of gas chromatography-ion mobility spectrometry to identify patients hospitalised with COVID-19 and predict prognosis

**DOI:** 10.1101/2022.02.28.22271571

**Authors:** Joshua Nazareth, Daniel Pan, Jee Whang Kim, Jack Leach, James G Brosnan, Adam Ahmed, Emma Brodrick, Alfian Wicaksono, Emma Daulton, Caroline Williams, Pranabashis Haldar, James A. Covington, Manish Pareek, Amandip Sahota

## Abstract

**Background:** Tests that can diagnose COVID-19 rapidly and predict prognosis would be significantly beneficial. We studied the ability of breath analysis using gas chromatography-ion mobility spectrometry (GC-IMS) for diagnosis of COVID-19 and as a predictor for subsequent requirement for Continuous Positive Airway Pressure (CPAP).

**Methods:** We undertook a single centre prospective observational study in patients with COVID-19, other respiratory tract infections and healthy controls. Participants provided one breath sample for GC-IMS analysis. We used cross validation analysis to create models that were then tested against the original cohort data. Further multivariable analysis was undertaken to adjust for differences between the comparator groups.

**Results:** Between 01/02/2021 and 24/05/2021 we recruited 113 participants, of whom 72 (64%) had COVID-19, 20 (18%) had another respiratory tract infection and 21 (19%) were healthy controls. Differentiation between patients with COVID-19 and healthy controls, and patients with COVID-19 and those with other respiratory tract infections, was achieved with high accuracy. Identification of patients with subsequent requirement for CPAP was completed with moderate accuracy and was not independently associated on multivariable analysis.

**Conclusions:** We have shown that GC-IMS has a high capability to distinguish between acute COVID-19 infection and other disease states. Breath analysis shows promise as a predictor of subsequent requirement for CPAP in hospitalised patients with COVID-19. This platform has considerable benefits due to the test being rapid, non-invasive and not requiring specialist laboratory processing.

## Introduction

Coronavirus disease (COVID-19) continues to cause significant global morbidity and mortality. Rapid diagnosis and prognostic assessment of patients with COVID-19 is crucial to ensure that patients can be triaged and managed appropriately. Current diagnosis of COVID-19 is made by RT-PCR on nasopharyngeal sampling, combined with clinical symptoms.^1^ However, nasopharyngeal samples have to be processed by trained laboratory staff and may be negative by the time a patient has symptoms severe enough to present to hospital.^2^

Exhaled breath analysis is an emerging approach to respiratory infection diagnosis. Volatile organic compounds (VOCs) measured in breath mirror metabolic processes both locally within the respiratory system, and systemically. Techniques to measure VOCs for diagnosis of infection have the potential to be rapid, non-invasive, at point-of-care, and completed without the need for trained laboratory staff.^3^ Breath analysis has already been shown to diagnose respiratory infections such as influenza, bacterial and tuberculosis with high accuracy.^3,4^ There is emerging evidence regarding the value of breath analysis in the diagnosis of patients with COVID-19 – however, evidence is still lacking regarding its ability to predict prognosis.

We therefore conducted a study to assess the ability of Gas Chromatography – Ion Mobility Spectrometry (GC-IMS) to identify hospitalised patients with COVID-19 from both healthy controls and patients with other respiratory infections. We also assessed whether GC-IMS was able to predict prognosis in hospitalised patients with COVID-19.

## Method

### Study settings

We undertook a prospective observational study between 01/02/2021 and 24/05/2021, which enrolled consecutive patients hospitalised for COVID-19, other infective respiratory tract infections, and healthy controls at Glenfield Hospital, University Hospitals of Leicester NHS Trust, Leicester, UK. During this period, there was a transition of SARS-CoV-2 variants in the UK from alpha to delta (98% of sequenced SARS-CoV-2 samples were classified as alpha or delta variants after May 10, 2021), and vaccinations were prioritised in the general population for older persons or those at risk of developing severe disease.^5^ Participants provided one breath sample for the GC-IMS instrument within 24 hours of a RT-PCR nasopharyngeal swab for SARS-COV-2.

For COVID-19, we included patients who fulfilled the following criteria: age≥ 16 years, hospitalised, tested positive for SARS-CoV-2 on RT-PCR using routine nasopharyngeal testing, no previously known positive SARS-CoV-2 RT-PCR or clinically diagnosed COVID-19 prior to the current admission. For respiratory tract infections, we recruited patients: aged≥ 16 years, hospitalised, tested negative for SARS-CoV-2 on RT-PCR using routine nasopharyngeal testing, with a clinical, radiological or microbiological diagnosis of another respiratory tract infection. Healthy controls were: aged ≥ 16 years, with no respiratory symptoms and no positive test for SARS-CoV-2 on RT-PCR in the preceding eight weeks before recruitment. Patients who were unable to understand and comply with the protocol, or unable or unwilling to give informed consent, were not included in the study.

### Clinical data collection

We collected demographic and clinical data on age, gender, ethnicity, comorbidities (autoimmune disease, hypertension, diabetes, ischaemic heart disease, chronic kidney disease, cancer, chronic lung disease, neurological disease, gastroenterological/liver disease, haematological disease), COVID-19 vaccination status, clinical symptoms at the time of sampling, duration between symptom onset and recruitment, radiology and laboratory findings (white cell count, lymphocyte count, and haemoglobin concentration as well as plasma concentrations of sodium, potassium, urea, creatinine, and C-reactive protein (CRP)) on admission. Laboratory findings were tested for on the same day as breath sampling. Clinical outcomes collected included requirements for non-invasive and invasive ventilation and death by 30^th^ June 2021.

### Ethics

The study had ethical approval from the West Midlands Research Ethics Committee (REC Reference 20/WM/0153). It was conducted in accordance with ICH-GCP, Declaration of Helsinki and the Data Protection Act 1998 and NHS Act 2006. All participants gave written, informed consent prior to any study procedures.

### Sampling platform

For this study a commercial GC-IMS instrument was used (G.A.S. BreathSpec™). This instrument has a relatively small footprint and is able to sit on a standard clinical trolley. It only requires a standard mains power supply and uses filtered room air as the carrier gas, provided by a circular gas flow unit (CGFU) fitted to the top of the unit. It comprises of a gas chromatograph front end for chemical separation, followed by a drift tube ion mobility spectrometer as the detector, providing dual separation of breath chemical components.^6,7^ A single breath sample was collected into a sterile 10ml syringe by aspirating from a specifically designed breathing apparatus at the time of exhalation. The syringe was then sealed in a bag and immediately taken to the BreathSpec™ machine for injection and processing (see supplementary pictures 1 & 2). Although possible to transport the machine to the patient, for infection control purposes the machine was located in a separate room to inpatients on the acute medical wards at Glenfield Hospital, Leicester.

### Statistical analysis

Continuous variables are expressed as median and interquartile range (IQR). Categorical variables are displayed as numbers and percentages (%). Pearson’s Chi-squared and Fisher’s exact row test was used to compare categorical variables between groups. Student’s *t-*test and Kruskal-Wallis test were used to compare continuous variables between groups depending on the normality of distribution. Analyses were performed using STATA version 14.2 (StataCorp United States) and Excel version 2016 (Microsoft, Redmond, United States).

The GC-IMS data was processed in line with the in-house pipeline developed at Warwick University.^6,7^ In brief, the data was pre-processed to reduce dimensionality and then a 10-fold cross validation was applied. Within each fold discriminatory features were identified by a rank-sum test and these features used to construct models (specifically Gaussian Process and Neural network), which was applied to the test set. This was repeated until all the data has been a test set and the resultant probabilities used to calculate statistical values.

Multivariable logistic regression analysis was used to investigate the relation between variables and the ability of GC-IMS to distinguish COVID-19 from other respiratory infections, as well as predict the prospective requirement of patients with COVID-19 for CPAP. Models adjusted for variables which were found to be different between groups on univariable analysis. A *p-* value <0.05 was considered to be statistically significant.

## Results

### Participant demographics and clinical features

Between February and June 2021, a total of 113 participants were recruited into the study; 72 were admitted to hospital with COVID-19; 20 were admitted to hospital with a respiratory tract infection other than COVID-19 and 21 were healthy controls (see Figure 1).

**Figure 1:**
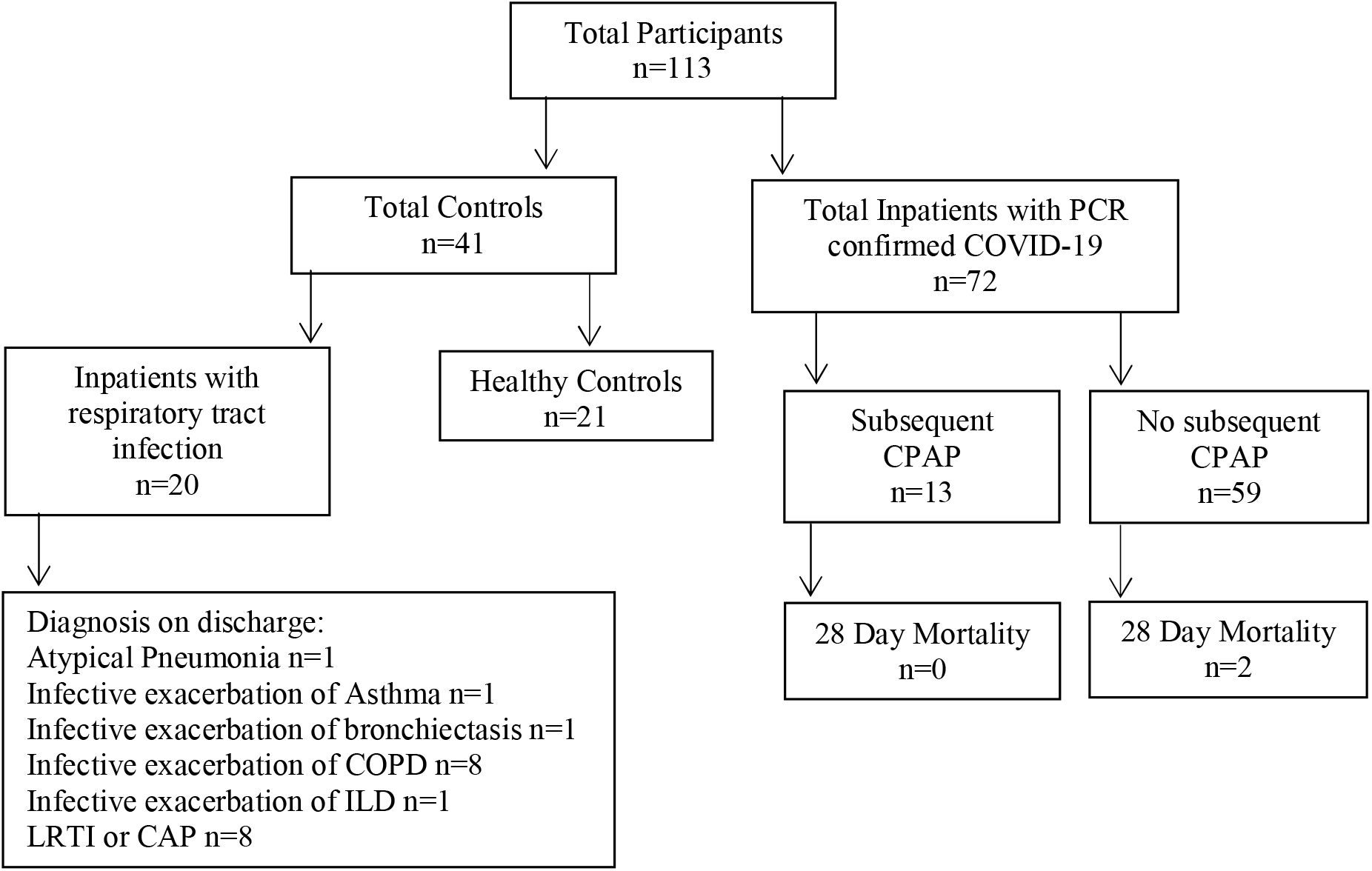
Details of participants recruited into the study. LRTI, Lower respiratory tract infection; CAP, Community acquired pneumonia; COPD, Chronic obstructive pulmonary disease; ILD, Interstitial lung disease

Table 1 shows participant demographic and clinical data. All respiratory controls had two PCR negative swabs for SARS-CoV-2. Twelve patients had radiological features on Chest imagine suggestive of infection; two had a microbiological diagnosis of infection either by respiratory PCR, sputum culture or blood culture; three had both a radiological and microbiological diagnosis of respiratory tract infection and three respiratory control patients had no positive microbiology/virology results or radiological changes and were diagnosed by a respiratory physician based on clinical features. Healthy controls were younger, more likely to be female and of White ethnicity compared to those who were admitted to hospital with a respiratory infection. Patients with COVID-19 were more likely to have lower white cell counts compared to other respiratory infections. Around a quarter of patients (27%) with COVID-19 had one dose of either the Oxford Aztrazeneca or Pfizer BioNTech vaccines prior to hospitalisation. Thirteen patients (18%) with COVID-19 required CPAP. There were four deaths in the 28 days following recruitment, two COVID-19 patients and two respiratory control patients.

**Table 1:** Participant demographic, clinical, laboratory and clinical outcomes.

| Variables | COVID +ve<br>n= 72 | Other respiratory<br>infection<br>n= 20 | Healthy<br>Controls<br>n= 21 | p value |
| --- | --- | --- | --- | --- |
| **Demographic data** |  |  |  |  |
| Age – median years (IQR) | 57 (49-66) | 65 (52-77) | 45 (38-51) |  |
| Male – n (%) | 47 (66) | 12 (60) | 3 (14) | <0.001 |
| White ethnicity – n (%) | 46 (75) | 18 (95) | 16 (76) | 0.004 |
| Asian ethnicity – n (%) | 14 (23) | 1 (5) | 2 (10) |  |
| Black ethnicity - n (%) | 1 (2) | 0 | 3 (14) |  |
| **Clinical data** |  |  |  |  |
| Autoimmune disease | 13 (18) | 6 (30) |  | 0.24 |
| Hypertension | 21 (29) | 7 (35) |  | 0.61 |
| Diabetes | 16 (22) | 4 (20) |  | 0.99 |
| Ischaemic heart disease | 12 (17) | 5(25) |  | 0.40 |
| Chronic kidney disease | 3 (4) | 3 (15) |  | 0.11 |
| Cancer | 1 (1) | 0 (0) |  | 0.99 |
| Chronic lung disease | 13 (18) | 15 (75) |  | <0.001 |
| Neurological disease | 2 (3) | 3 (15) |  | 0.07 |
| Gastroenterological/liver disease | 7 (10) | 1 (5) |  | 0.68 |
| Haematological | 4 (6) | 1 (5) |  | 0.99 |
| Number of comorbidities | 1.5 (0-2) | 2.5 (1-3) |  | 0.04 |
| Admission oxygen saturations – median % (IQR) | 97 (95-98) | 96 (94-97.5) |  | 0.35 |
| White cell count – median x10<sup>9</sup> cells/L (IQR) | 7.1 (5.3-9.1) | 11.8 (10.3-14.0) |  | <0.001 |
| Lymphocyte - median x10<sup>9</sup> cells/L (IQR) | 1.0 (0.7-1.5) | 1.1 (0.9-2.3) |  | 0.34 |
| Urea – median mmol/L (IQR) | 5.3 (3.9-7.2) | 5.7 (4.2-9.4) |  | 0.56 |
| Creatinine – median μmol/L (IQR) | 82.5 (66.5-99) | 87.5 (68.5-115) |  | 0.15 |
| CRP – median mg/L (IQR) | 80.5 (35-138.5) | 51 (13.5-140.5) |  | 0.59 |
| Haemoglobin – median g/L (IQR) | 142.5 (131.5-153) | 124 (116.5-146) |  | 0.06 |
| Duration of symptoms – median days (IQR) | 7 (5-10) | 7 (2.5-8) |  | 0.51 |
| Treatment with dexamethasone – n (%) | 55 (78) |  |  |  |
| Vaccinated - n (%) | 19 (27) |  |  |  |
| Pfizer – n (%) | 12 (17) |  |  |  |
| Astra Zeneca – n (%) | 7 (10) |  |  |  |
| **Clinical outcomes** |  |  |  |  |
| Need for supplemental oxygen during hospital admission – n (%) | 57 (79) | 14 (70) |  | 0.39 |
| Received CPAP following sampling – n (%) | 13 (18) |  |  |  |
| 28 day mortality – n (%) | 2 (3) | 2 (10) |  |  |

^ t-test comparing differences in age between COVID +ve and Other respiratory infection groups only 11 participants had missing ethnicity data, there was no other missing data

Table 2 shows demographics of patients with COVID-19, stratified by whether they had received CPAP during hospitalisation. Participants that required CPAP following sampling were more likely to have higher admission serum urea levels and a longer duration of symptoms prior to sampling compared to those who did not.

**Table 2:**
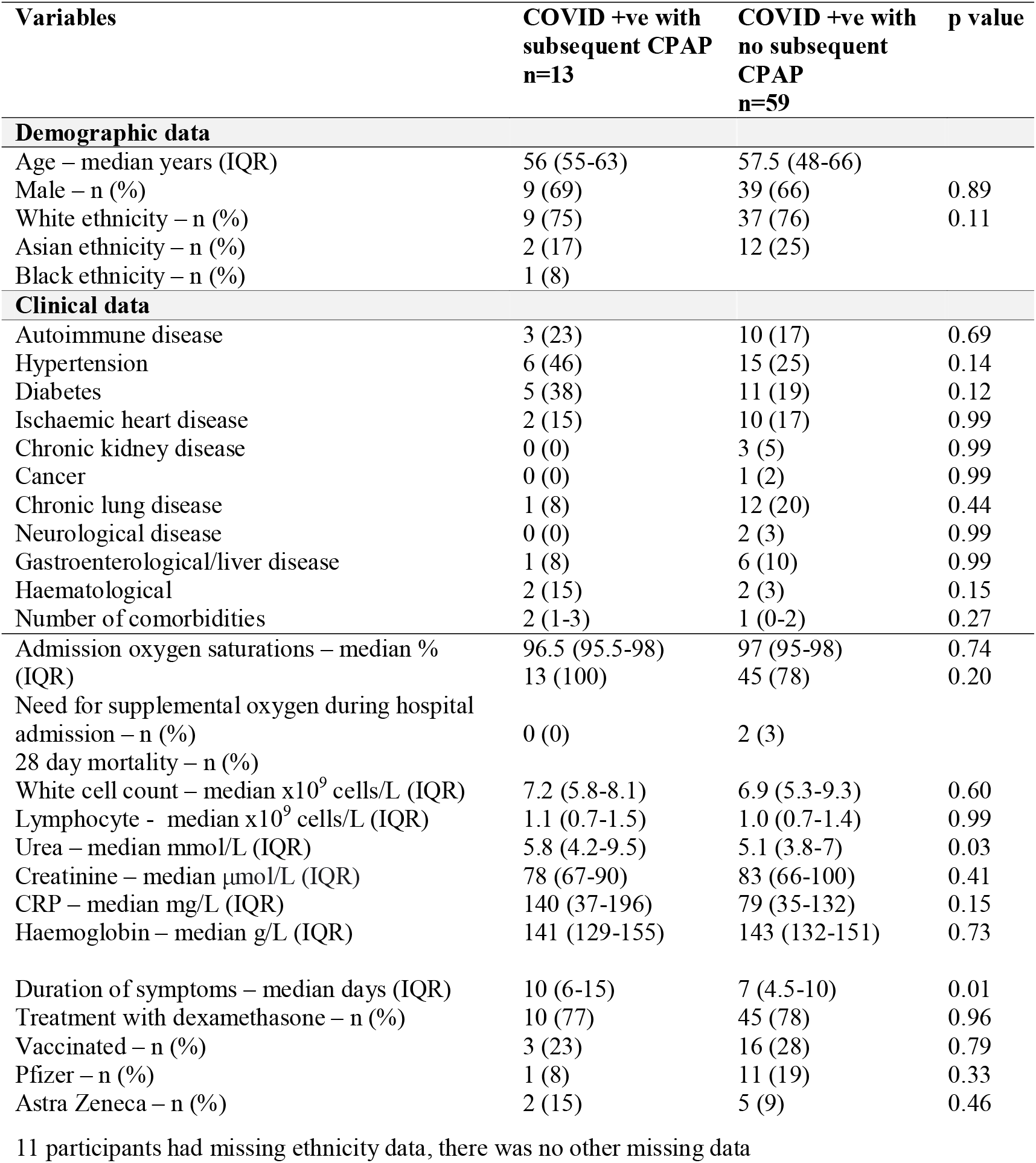
**COVID-19 positive Participant demographic, clinical, laboratory and clinical outcomes by future requirement for CPAP**

### GC-IMS results

Table 3a shows the results from GC-IMS instrument, comparing performance metrics for the ability of the instrument to distinguish between COVID-19 and healthy controls, COVID-19 and other respiratory infections, as well as COVID-19 disease severity (as defined by requirement for CPAP). Overall the machine was able to distinguish between all three states with highly significant *p* values across all groups.

**Table 3:** **Performance metrics for the three comparisons made: COVID-19 vs Healthy Control, COVID-19 vs Respiratory control, COVID-19 that subsequently required CPAP vs COVID-19 with no subsequent requirement for CPAP**

|  | COVID-19 vs Healthy Control | COVID-19 vs Respiratory Control | COVID-19 +ve Subsequent CPAP vs No subsequent CPAP |
| --- | --- | --- | --- |
| Classifier | Neural Network | Gaussian Process | Neural Network |
| AUC (95%ci) | 0.85 (0.74-0.96) | 0.89 (0.81-0.96) | 0.70 (0.53-0.87) |
| Sensitivity (95%ci) | 0.80 (0.56-0.94) | 0.85 (0.76-0.91) | 0.62 (0.32-0.86) |
| Specificity (95%ci) | 0.88 (0.79-0.94) | 0.90 (0.68-0.99) | 0.80 (0.69-0.89) |
| Positive Predictive Value | 0.59 | 0.98 | 0.36 |
| Negative Predictive Value | 0.95 | 0.56 | 0.92 |
| p value | <0.001 | <0.001 | 0.01 |
| Threshold | 0.06 | 0.66 | 0.06 |
| Multivariable analysis ^ | Adjusted OR (95% CI) | P value |  |
| Outcome: COVID-19 vs Respiratory Control |  |  |  |
| Breath analysis Gaussian Process* | 2.35 (1.55-3.57) | <0.001 |  |
| Admission White cell count | 0.73 (0.58-0.91) | 0.01 |  |
| Number of comorbidities | 0.97 (0.56-1.69) | 0.92 |  |
| Outcome: Subsequent CPAP vs no subsequent CPAP |  |  |  |
| Breath analysis Neural Network† | 1.13 (0.95-1.35) | 0.17 |  |
| Admission Urea | 1.04 (0.93-1.17) | 0.47 |  |
| Duration of symptoms<sup>Δ</sup> | 1.15 (1.00-1.33) | 0.047 |  |

OR, Odds ration; CI, Confidence interval; ^ Analyses adjusted for all other variables in the table.
\*Gaussian Process COVID +ve Vs Respiratory control analysis output was multiplied by 10 in order for multivariable regression to be completed;
†Neural network Subsequent CPAP vs No subsequent CPAP analysis output was multiplied by 10 in order for multivariable regression to be completed;
<sup>Δ</sup>for each day increase in duration of symptoms

Different classifiers generated the best distinguishing ability between groups, with Neural Network being the best to distinguish between COVID-19 and healthy controls; Gaussian Process to distinguish between COVID-19 and other respiratory infections, and Neural Network for CPAP vs no CPAP. Using these classifiers, we found GC-IMS to have a positive predictive value of 98% in distinguishing between COVID-19 or another respiratory infection in hospitalised patients, a negative predictive value of 97% in distinguishing between COVID-19 vs a healthy control, and a negative predictive value of 92% in identifying subsequent requirement for CPAP in hospitalised COVID-19 patients. ROC curves using different classifiers for all analyses are shown in Supplementary Materials.

### Multivariable analysis of predictors of correct identification of patients with COVID-19 and subsequent requirement for CPAP

Table 3b shows adjusted logistic regression analyses of GC-IMS and other variables in relation to prediction of COVID-19 compared to other respiratory infections, and prospective requirement for CPAP. In an adjusted analysis, both decreasing serum white cell count and increasing GC-IMS readings (using Gaussian Process) were independently related to a diagnosis of COVID-19 compared to other respiratory infections. However, longer duration of COVID-19 symptoms prior to admission to hospital but not increasing GC-IMS readings (using Neural Network) was independently associated with prediction of prospective requirement for CPAP.

## Discussion

There are three main findings from this study. Firstly, GC-IMS is feasible and breath collection by the selected method well tolerated in patients admitted to hospital with COVID-19. Secondly, GC-IMS was able to distinguish COVID-19 infection from other respiratory infections, as well as healthy controls. Thirdly, we found a relationship between GC-IMS readings and worse prognosis in COVID-19, as evidenced by an association with prospective requirement for CPAP.

We found exhaled breath analysis to be highly feasible in acutely unwell patients and that GC-IMS is strongly able to distinguish between COVID-19 from the other groups. Three previous studies have evaluated the diagnostic potential of breath analysis for COVID-19.^8–10^ All showed specific breath metabolomic signatures in patients with COVID-19 compared to controls with other diseases (respiratory infection, acute respiratory distress syndrome and chronic diseases which cause breathlessness), as well as healthy controls, with comparable receiver operating curves to those found in our study. In contrast to other studies, where patients had to be transported into a room housing the analysis machine, the BreathSpec™ machine we employed was portable and could be taken to the patients’ bedside. This would allow for use within routine hospital settings where patients may be too unwell to mobilise or transfer.

Our findings support the use of exhaled breath analysis in COVID-19 diagnosis. This has logistical advantages over current PCR testing, due to its rapidity in producing a result which can be performed and interpreted by non-laboratory staff.^11^ In hospital, this could allow for faster identification and triage of patients with COVID-19 from other respiratory infections, preventing nosocomial infection and faster commencement on appropriate therapy. Within primary care, application of such a test has the potential to distinguish bacterial from viral infections, helping clinicians to decide whether antibiotic prescriptions would be necessary as well as rapid direction of ill patients who have COVID-19 to hospital. Within other community settings e.g. schools and airports, such testing could offer significant benefits over current methods of rapid detection, and better tolerability to oropharyngeal swabbing.

To our knowledge, we are the first to demonstrate the potential of exhaled breath analysis to predict the need for requiring CPAP. Grassin-Delyle and colleagues found VOC concentrations were not correlated with severity of illness, as measured by concomitant severity scores (SAPS II and SOFA).^10^ However, one-off measurements of such scores underpredict disease severity in COVID-19, and the use of clinically important endpoints, such as CPAP may be more accurate as an outcome.^12,13^ Since only two participants died within our cohort, it remains unclear whether the differences we identified in exhaled breath metabolomics are a consequence of protective or deleterious immune responses within the lungs. Ruskiewicz and colleagues identified differences in exhaled breath metabolomics between those with mild COVID-19 compared to those who had fatal disease and those requiring intubation/intensive care, suggesting the latter hypothesis. In contrast to previous studies, we showed that GC-IMS has the potential to detect COVID-19 and predict disease trajectory in those who had been partially vaccinated. Thirdly, we did not perform genetic sequencing to examine for any differences in exhaled volatile compounds between different variants – however, this was not the intention of our study and it is likely that our technology can detect disease in multiple variants given the epidemiological transition from alpha to delta at the time.^5^ Finally, despite univariable associations, we did not find GC-IMS to be associated with subsequent CPAP requirement on multivariable analysis. Given that COVID-19 in its most severe disease states results in multi-organ failure, it may be that simply sampling from the respiratory tract is insufficient to provide the full clinical picture or the instrument does not have sufficient sensitivity. We note that a longer duration of symptoms prior to hospital admission was an independent predictor of prognosis - therefore GC-IMS may have stronger diagnostic and predictive roles in the clinical assessment of COVID-19 in early disease.

Our study was limited by sample size. Not all participants who had respiratory infections other than COVID-19 had positive PCR results or microbiology. Since nasopharyngeal tests for SARS-CoV-2 have limited sensitivity, it is therefore possible that our control groups’ symptoms could be explained by SARS-CoV-2 infection. However, no significant differences existed in the duration of symptoms between those that had COVID-19 and those with other respiratory infections, which is the main factor in relation to PCR positivity for COVID-19, with those early in infection most likely to have a positive test. Our study population was highly diverse, comprising of multiple comorbidities, vaccinations and treatment within hospital, which could have resulted in underestimation of GC-IMS’ ability to distinguish between specific subgroups.

In conclusion, GC-IMS has a high capability to distinguish between acute COVID-19 infection and other disease states, including other respiratory infections. GC-IMS also shows the ability to predict subsequent requirement for CPAP in hospitalised patients with COVID-19, but is not an independent predictor of outcome when other variables are taken into account. Our study demonstrates the use of a novel technology that can be embedded immediately within clinical practise, with workforce and economic implications that come with a reduced need of laboratory processing.

## Supporting information

Supplemental Figures and Pictures

## Data Availability

All data relevant to the study are included in the article or uploaded as supplementary information

## Declarations

### Funding

This study did not receive any funding

### Data Sharing

All data relevant to the study are included in the article or uploaded as supplementary information

## Notes

### Competing Interest Statement

The authors have declared no competing interest.

