## Supplemental Figures and Pictures for "Discriminatory ability of gas chromatography-ion mobility spectrometry to identify patients hospitalised with COVID-19 and predict prognosis"

**Supplementary Material**

**Supplementary figure 1**

Receiver Operator Characteristic curves for the two different classifiers used to distinguish between COVID-19 vs Healthy Control, COVID-19 vs Respiratory control, COVID-19 inpatients that subsequently required CPAP vs COVID-19 inpatients with no subsequent requirement for CPAP

COVID-19 vs Healthy Control

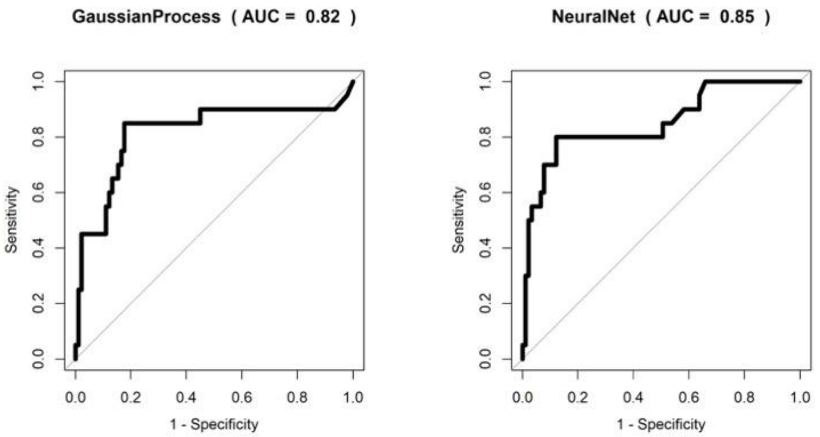


COVID-19 vs Respiratory control

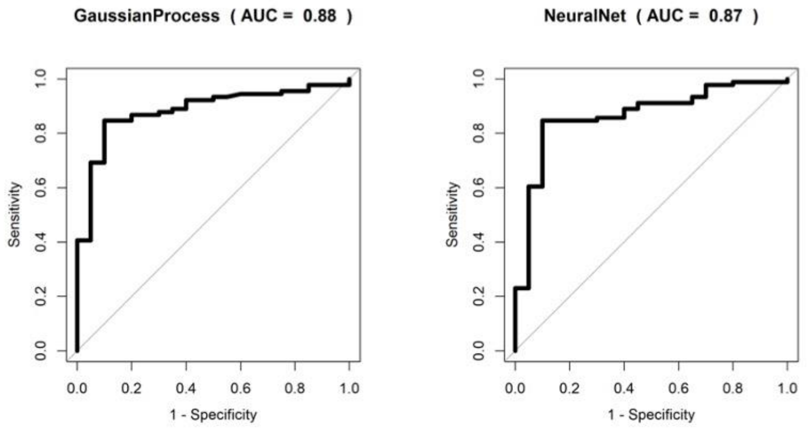


COVID-19 inpatients that subsequently required CPAP vs COVID-19 inpatients with no subsequent requirement for CPAP

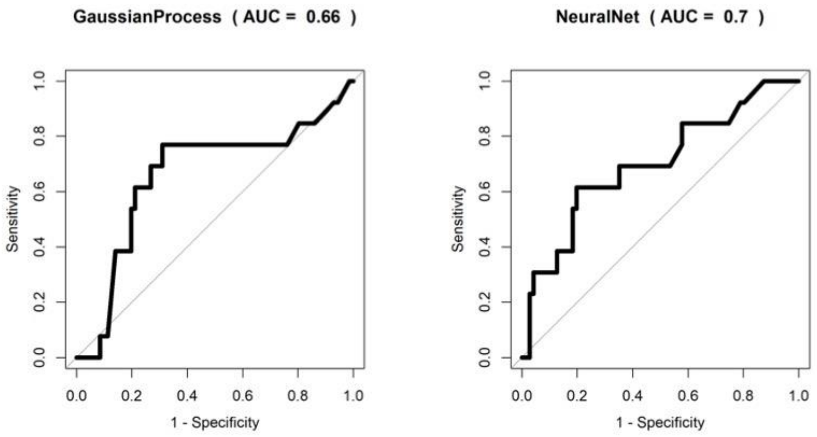

**Supplementary picture 1**
Collection of breath sample into 10ml syringe

*Image removed as it contains a photograph of a person. Please contact the corresponding author to request a copy of this picture.*

**Supplementary picture 2**
GC-IMS Breath analysis instrument (G.A.S. BreathSpec™)

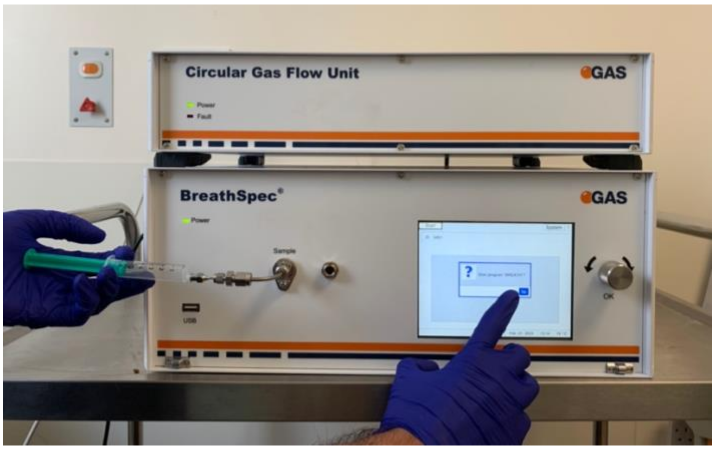
